# Modulation of Corticospinal Excitability and Muscle Synergies During Complex Locomotor Task in Individuals With and Without Cerebral Palsy: A TMS-EMG Study

**DOI:** 10.1101/2025.08.15.25331834

**Authors:** Yosra Cherni, Marco Ghislieri, Laurent Bouyer, Catherine Mercier

## Abstract

**Introduction:** Studies employing transcranial magnetic stimulation and electromyography suggest that disrupted functional corticospinal connectivity significantly contributes to locomotor impairments. In individuals with Cerebral Palsy (CP), the corticospinal tract has therefore been identified as a potential target for improving gait control. However, it remains uncertain whether this pathway can be further activated given the damage caused by the brain lesion. Moreover, muscle synergies, a cooperative activation of groups of muscles, play an essential role in efficient and adaptive locomotion. Understanding and improving the modulation of these synergies could lead to better rehabilitation strategies for individuals with CP. The objective of this study was to assess whether a complex walking task promotes an increase in corticospinal excitability and a modulation of muscle synergies compared to a simple walking task in individuals with CP.

**Methods:** Fourteen individuals with CP and fourteen control subjects took part in this study. Each participant performed a simple walking task and a complex walking task (i.e., stepping onto virtual targets) at comfortable speed, in counterbalanced order. Motor evoked potentials (MEPs) in the tibialis anterior muscle were induced using transcranial magnetic stimulation. Muscle synergies were extracted from surface electromyography signals acquired from six key lower-limb muscles during both tasks.

**Results:** MEPs were elicitable in 11/14 participants for the CP group and 14/14 for control group. In the complex task, MEPs increased by 59.4% in the CP group (MEP_simple task=1.89 [1.00-3.09] vs. MEP_complex task=2.70 [1.59-4.80] mV/s^2^; p≤0.01) and 113.8% in the control group (MEP_simple task=1.95 [0.99-2.72] vs. MEP_complex task =2.91 [1.97-3.66] mV/s^2;^ p≤0.01). An increase in the number of synergies was observed during complex task in CP group (p=0.018).

**Conclusions:** These results suggest that performing a complex walking task allows to enhance the corticospinal excitability in both individuals with CP and control subjects. Moreover, CP individuals showed that either the number or the structure of synergies are modulated by the complex task, in comparison to control subjects. Longitudinal studies are recommended to assess the impact of the integration of complex tasks in gait rehabilitation interventions.

## 1. Introduction

Cerebral palsy (CP) is the most common motor disability in childhood, often resulting from a defect or lesion in the developing brain(1). While the etiology of CP varies, damage to the corticospinal tract (CST), which connects the cerebral cortex to the spinal cord and plays a key role in voluntary motor control, is a common feature of CP (2,3). This damage affects the signals that are sent from the motor cortex to the muscles, leading to a range of motor deficits. Studies based on non-invasive brain stimulation techniques, such as transcranial magnetic stimulation (TMS), and on surface electromyography (sEMG) showed that individuals with CP often exhibit reduced corticospinal excitability (4,5). This means that the descending drive from the motor cortex to the muscles is weaker, leading to difficulties in initiating and controlling voluntary movements. This may be reflected in a reduction in performance in daily living tasks, including walking (6–8).

Muscle synergy theory has been recently proposed to quantitatively and non-invasively assess the motor control system both in physiological and pathological conditions (9,10). This theory suggests that instead of independently controlling each muscle, the central nervous system may coordinate the activation levels and timing of all muscles involved in a specific movement using a small number of neural commands. Specifically, the muscle synergy theory can be derived from sEMG data using specific factorization methods, enabling the study of underlying motor control patterns and strategies that are believed to reflect neural function (10). Even if the neural origin of muscle synergies is still a topic of discussion among the scientific community (11), several studies have demonstrated the usefulness of the muscle synergy theory in describing motor control system of patients with orthopedical and neurological diseases affecting movements (12– 15).

From this perspective, muscle synergies may represent a valuable tool for studying motor control alterations in CP patients who are expected to exhibit altered corticospinal excitability. In fact, altered corticospinal excitability is commonly associated with motor impairments, such as spasticity and weakness (16,17), and changes in this variable are believed to reflect neuroplasticity (18). Therefore, corticospinal excitability alterations may disrupt both the composition and the number of muscle synergies (19,20), leading to less coordinated and more stereotyped movements. In some cases, individuals may also rely on pathological movement patterns to compensate for the motor deficits. Human locomotion is a widely studied movement for assessing motor control. It is one of the most important activities of daily living and can be easily performed by patients who can walk independently without aid or external support. In daily-life activities, humans often must adapt their gait to avoid obstacles or speed changes. Unpredictable disturbances, such as any deviation from regular walking, can lead to falls and injuries (21,22). Much of the existing research has focused on the automatic control of walking by studying repetitive movements in highly controlled conditions (e.g., walking on a straight-line path or treadmill). These “simple” walking tasks predominantly engage automatic neural circuits^3^, like the spinal structures, and depend less on voluntary control circuits, such as the CST (23), which are crucial for daily-life adaptations. Measuring corticospinal excitability and sEMG data during walking conditions mainly involving voluntary control circuits (i.e., the corticospinal tract) may provide insights into how the CNS controls lower-limb muscles in a more ecological environment. This is crucial for understanding the neural mechanisms of gait, which is a complex and fundamental task of daily living. In addition, although it is commonly accepted that muscle synergies are controlled at the spinal cord level(24), cortical contributions may be essential for ensuring a flexible and goal-directed motor control. In control subjects, evidence suggests that fundamental muscle synergy structures (synergy weights or vectors) remain consistent despite external environmental changes during walking (25,26), such as varying gait speeds (27,28), different slopes (29), or distinct walking tasks (e.g., nordic or beam walking) (30,31). However, in children with CP, Kim et al. (32) showed that muscle synergies are more sensitive to changes during walking tasks (e.g., self-selected speed, variable speed, and restricted width) than in typically developing children. Investigating both corticospinal excitability and muscle synergies during regular and complex walking tasks may improve our knowledge about locomotor control in individuals with CP and refine rehabilitation intervention.

The objectives of this study were to: (1) assess whether a complex walking task promotes an increase in corticospinal excitability and a modulation of muscle synergies compared to simple walking task in individuals with CP and controls and (2) to evaluate whether the change is different between the two groups. We hypothesized that an increase in the corticospinal excitability (i.e., complex task) might further involve the corticospinal tract and, thus, determine a reorganization of the locomotor muscle synergies (in terms of number and robustness of the muscle synergies). Moreover, it was expected that control subjects would show a greater increase in corticospinal excitability compared to individuals with CP. In terms of muscular synergy, we expected a modulation of the number and structures.

### 2. Materials and Methods

### 2.1. Participants

CP and control groups were recruited for this study. For both groups, the inclusion criteria were:

(1) to be aged between 12 and 45 years; (2) to be able to walk without walking aid for approximately 3 minutes. To be eligible to perform the corticospinal excitability assessment part of the study, participants must not have any exclusion criteria for TMS as detailed by Rossi et al. (33). Inclusion criteria specific to the CP group required participants to have a confirmed diagnosis of unilateral or bilateral cerebral palsy and an ability to understand simple instructions (e.g., walks normally, walks on targets, etc). For both groups, the exclusion criteria were any unstable health condition that could affect task performance. Participants with contraindications to TMS performed part of the protocol, which involved only sEMG measurements without TMS. Prior to taking part in the study, all participants provided their informed consent for the experimental procedure in accordance with the Declaration of Helsinki. The study was approved by the Quebec City Rehabilitation Institute Ethics Review Board (2021-2209, RIS).

### 2.2. Experimental protocol

Individuals with CP eligible for TMS were assessed during two evaluation sessions. The first session involved Magnetic Resonance Imaging (MRI) to obtain the patient-specific brain anatomical structure, which was then used to guide the experimenter in positioning the TMS coil. During the second session, CP patients underwent neurophysiological measurements (i.e., motor evoked potential (MEP) and sEMG) while performing two consecutive treadmill walking tasks at a comfortable self-selected speed: *a*) a Simple Walking (SW) task and *b*) a Complex Walking (CW) task. As developed in our previous study (34), the CW task involves walking on a treadmill and placing the feet on virtual targets presented at different intervals (80%, 100%, 120% of the normal step length) on a screen facing the subject. The real-time positions of the feet were also displayed on the screen as feedback, with each foot represented by a small sphere centered at the heel position. The distance between visual targets in the sagittal plane randomly varied with each step. During the SW task, instead, no visual targets were displayed on the screen facing the participants. Control participants attended only the second evaluation session (i.e., neurophysiological measurements), a template brain being used for the positioning of the TMS coil.

### 2.3. Data acquisitions

#### Corticospinal excitability

MEPs from Tibialis Anterior (TA) muscle were assessed using single-pulse TMS (Magstim Rapid2; custom “batwing” coil (34,35)) applied to the motor cortex (contralateral to the most**-**affected lower limb in CP subjects and the non-dominant lower limb in control subjects). The hotspot and active motor threshold were determined while the participant was in a seated position, with knees flexed at 90 degrees and maintaining approximately 20 degrees of dorsiflexion against gravity to facilitate MEPs. The hotspot was identified as the zone that evokes MEPs at the lowest stimulation intensity (36). The active motor threshold (aMT) was determined at the hotspot as the lowest stimulation intensity required to evoke a MEP in 5/10 trials (37). Then, single-pulse stimulations at 110% of the aMT were administered during walking, with 25 stimulations per condition. These stimulations occurred around 40% of the walking cycle, specifically in the late stance phase when the activity of the TA muscle is minimal, to ensure consistency in comparisons across different conditions (34). The timing corresponding to 40% of the gait cycle was determined in real time using the treadmill force plates. A neuronavigation system (Brainsight, Rogue research, Montreal, Canada) guided the experimenter while positioning the coil above the experimentally determined hotspot. The position of the coil at the moment of application of each pulse was recorded to allow offline rejection of off-target stimuli and comparison of coil positioning across conditions. Additionally, the position of each foot was tracked using 3 photo-reflective markers and visualized in real-time using an optoelectronic system equipped with 8 infrared cameras (Vicon Motion Systems). To avoid patrticipants’ anticipation, stimulations were provided on random gait cycles. On average, a single pulse stimulation was provided every 5 strides with a range of 3-7 strides. Participants not eligible for TMS took part only in the protocol assessing muscle synergies, described in the following section.

#### Muscle activity

sEMG signals of the most affected (CP group) or non-dominant (control group) lower limb were recorded by focusing on the muscles involved in the walking tasks. In particular, the following 6 lower-limb muscles were monitored: TA, Soleus (SOL), Gastrocnemius lateralis (GL), Rectus femoris (RF), Vastus lateralis (VL), and Semitendinosus (ST). The ground electrode was positioned on the patella. Ag/AgCl electrodes (Kendall Medi-trace 200, Covidien) were placed on the volunteers’ skin over the belly of each muscle. Prior to electrode placement, the skin was shaved and cleaned to minimize the impedance between the skin and the electrode. Each electrode location was determined according to the recommendations of Surface EMG for Non-Invasive Assessment of Muscles (SENIAM) (38). The ground electrode was positioned on the patella of the homolateral lower limb. All signals were sampled at 1000 Hz.

### 2.4. Data analysis

#### 2.4.1. MEP analysis

To quantify corticospinal excitability, the area under the curve was calculated for each rectified MEP and then averaged for each task and participant(34). In each group, the CST excitability change between the two tested conditions (i.e., MEP modulation) was measured as follows:

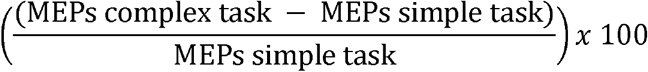

The error in TMS coil location was measured through the neuronavigation software as the distance between the location of each stimulation and the hotspot (expressed in millimeters). Finally, the mean background sEMG amplitude was measured (considering 50-ms windows preceding every TMS stimulation) and compared across task conditions. All data were analyzed offline by using custom MATLAB routines (MATLAB r2018a, The MathWorks Inc., Natick, MA, USA).

#### 2.4.2. Muscle synergy extraction

Raw sEMG signals were band-pass filtered between 10 Hz and 450 Hz by means of a zero-lag 4th-order Butterworth filter(39). Thereafter, filtered sEMG signals were full-wave rectified and the sEMG envelopes were obtained using a 4-th order low-pass Butterworth filter with a cut-off frequency set at 10 Hz (40). Then, separately for each muscle, the resulting envelopes were amplitude-normalized by the maximum of each sEMG envelope obtained during the SW task. A 5% body weight vertical cut-off force threshold was applied to the ground reaction forces to identify gait events. Gait cycles duration was normalized to 1000 samples to avoid biases due to different gait cycle durations.

The Non-Negative Matrix Factorization (NNMF) was used to extract locomotor muscle synergies during both walking conditions(41–43). The NNMF algorithm involves an iterative factorization of the sEMG envelope matrix *E*, which has dimensions *t* × *m*. It decomposes *E* into two matrices: the activation coefficient matrix *C* with dimensions *t* × *m* and the weight vector *W* with dimensions *s* × *m* This can be expressed as follows:

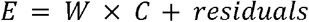

The iterations continue until the Frobenius norm of the residuals is minimized. In this context, *t* represents the number of time instants, *m* represents the number of muscles, and *s* represents the final number of muscle synergies. For further details, please refer to Hug et al.(44).

In the present study, matrix *E* contains the time- and amplitude-normalized sEMG envelopes It has dimensions (*N* × *t*) × *m*, with *N* denoting the number of gait cycles, *t* representing the number of time instants (*t* = 1000), and representing the number of acquired muscles (*m* = 6). Different NNMF solutions were tested by running several times the algorithm on the same sEMG data changing the muscle synergy number (*s*) between 1 and 6 (i.e., the total number of muscles acquired). For each number of muscle synergies, the reconstruction quality was assessed using the Variance Accounted For (VAF) parameter. Consequently, the final number of muscle synergies required to properly reconstruct the original sEMG signals was defined by choosing the smallest number of synergies granting a VAF value equal or greater than 90% (45). Muscle synergies were extracted through custom MATLAB routines (MATLAB r2018a, The MathWorks Inc., Natick, MA, USA).

### 2.5. Statistical analysis

**For the groups’ characteristics,** parameter estimates are represented as median and interquartile. In all the analyses, the significance level (α) was set to 0.05. Mann-Whitney U-tests were used to test differences in anthropometric characteristics between groups (i.e., CP and control).

**To test our first objective related to corticospinal excitability**, the nparLD test (46), a non-parametric test equivalent to a repeated-measures ANOVA, was applied to MEP results (i.e., MEP under the curve area, EMG background and coil error), with the within-subject factor *Condition* (i.e., SW and CW tasks) and the between-subject factor *Group* (i.e., CP and control). In the presence of a significant interaction, *post-hoc* analyses were carried out. NparLD analysis was carried out using RStudio software. Between groups’ MEP modulations were compared using a Mann-Whitney U test.

**To test our second objective related to muscle synergies**, a Wilcoxon test was used to compare the final number of muscle synergies between SW and CW tasks in CP subjects. Note that there is no group factor in this analysis as the control group served as a reference. The number of synergies was determined for each condition in the control group. Specifically, the N synergies identified in the control group during the SW were used as a reference for comparison with the synergies of the CP group performing the same task. Similarly, the N synergies identified in the control group during the CW served as a reference for comparison with the synergies of the CP group in that task. To assess differences in motor control strategies between individuals with CP and control participants, the average activation coefficients 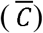 and weight vectors 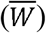 extracted during the walking tasks of control individuals were used as a reference. Then, the Cosine Similarity (*S*) was used to assess the similarity between the muscle synergies of each CP individual and the reference (i.e., muscle synergies averaged over control group) considering both *C* and *W* vectors. The *S* value may range between 0 (totally dissimilar) and 1 (totally similar)(47). Muscle synergies characterized by a *CS* value below 0.8 were classified as *subject-specific* muscle synergies, whereas muscle synergies characterized by a *S* value equal to or higher than 0.8 were classified as *shared* (or *common*) muscle synergies. Muscle synergies were compared using custom MATLAB scripts.

## 3. Results

### 3.1 Participants

Fourteen individuals with CP and fourteen control subjects were included in the present study. Groups’ characteristics are reported in **Table 1**. Individuals with CP had Gross Motor Function Classification System (48) (GMFCS) scores of I (n=3), II (n=8) and III (n=3). Three participants had contraindication to TMS due to a history of epilepsy. Consequently, corticospinal excitability measurements were conducted on 11 CP patients and 14 healthy controls. The characteristics of the CP individuals and controls included in the study are detailed in **Table 1**.

**Table 1.** Characteristics of CP and control groups.

|  | Sex | Age<br>(years) | Mass<br>(kg) | Heigh<br>(cm) | aMT<br>(%) |
| --- | --- | --- | --- | --- | --- |
| CP (n=14) | 5♀ | 27 [16-35] | 63 [56-80] | 166 [162-175] | 65 [64-70]* |
| CTL (n = 14) | 8♀ | 25 [23-28] | 64 [58-79] | 170 [165-178] | 59 [55-64] |
| p-value | 0.276 | 0.322 | 0.662 | 0.433 | **0.005** |

Parameter estimates are represented as mean ± standard deviation over the group. P-values represent the results of the comparison between groups using Mann-Whitney U-tests. CP: Cerebral Palsy participants; CTL: Control participants; aMT: active motor threshold; \*only 11/14 CP participants had TMS assessments. Significant statistical differences are highlighted in ***italic bold***.

### 3.2. Corticospinal excitability

Figure 1. shows results related to corticospinal excitability (i.e., MEP under curve area and MEP modulation), presented as median and interquartile range across each group. The NparLD results showed a significant effect of the condition (SW *vs.* CW; p < 0.0001), but not of the group (CP *vs* Control; p=0.905) nor a condition x group interaction (p=0.332). Based on the Mann-Whitney U test, MEP modulation did not differ significantly between groups (p=0.396, Figure 2B). An effect of the condition was observed in the sEMG background (p=0.015), without any effect of the group or condition x group interaction (p=0.691 and 0.626, respectively). Regarding the coil placement during walking tasks, the target errors of the included stimulation were less than 5 mm and no significant effects of group, condition, or group x condition interaction were observed (p>0.05).

**Figure 1.**
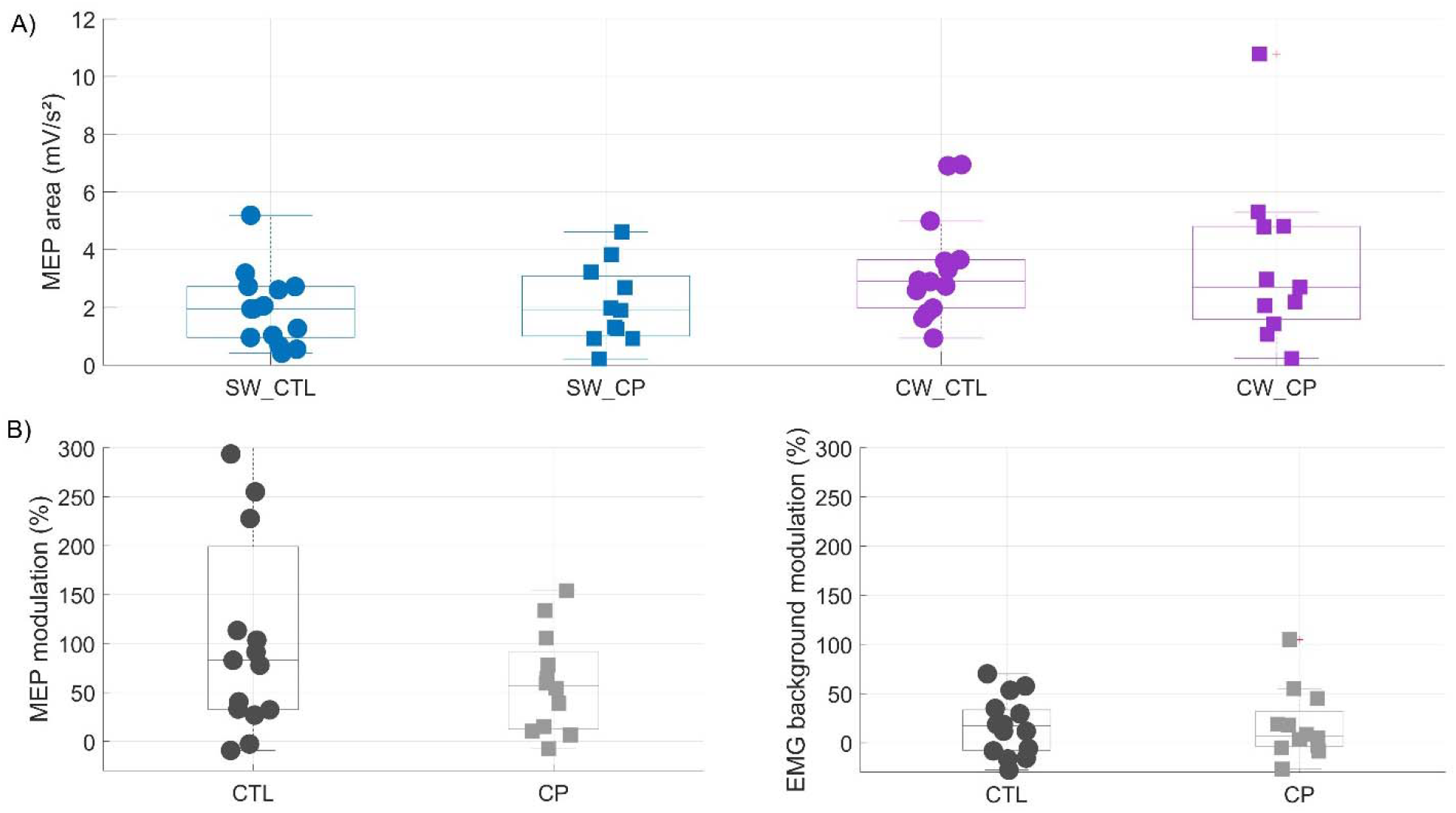
(A) Corticospinal excitability during Simple Walking (SW, blue) and Complex Walking (CW, purple) tasks in cerebral palsy (CP, ◼) and Control (CTL, ◼) groups. NparLD p-value for the condition effect (SW vs. CW) is displayed. No significant effect for the group factor (CP vs Control) nor at the interaction level (condition x group) are observed. (B) Modulation (%) between the two tasks in terms of motor evoked potential (MEP) and EMG background.

**Figure 2.**
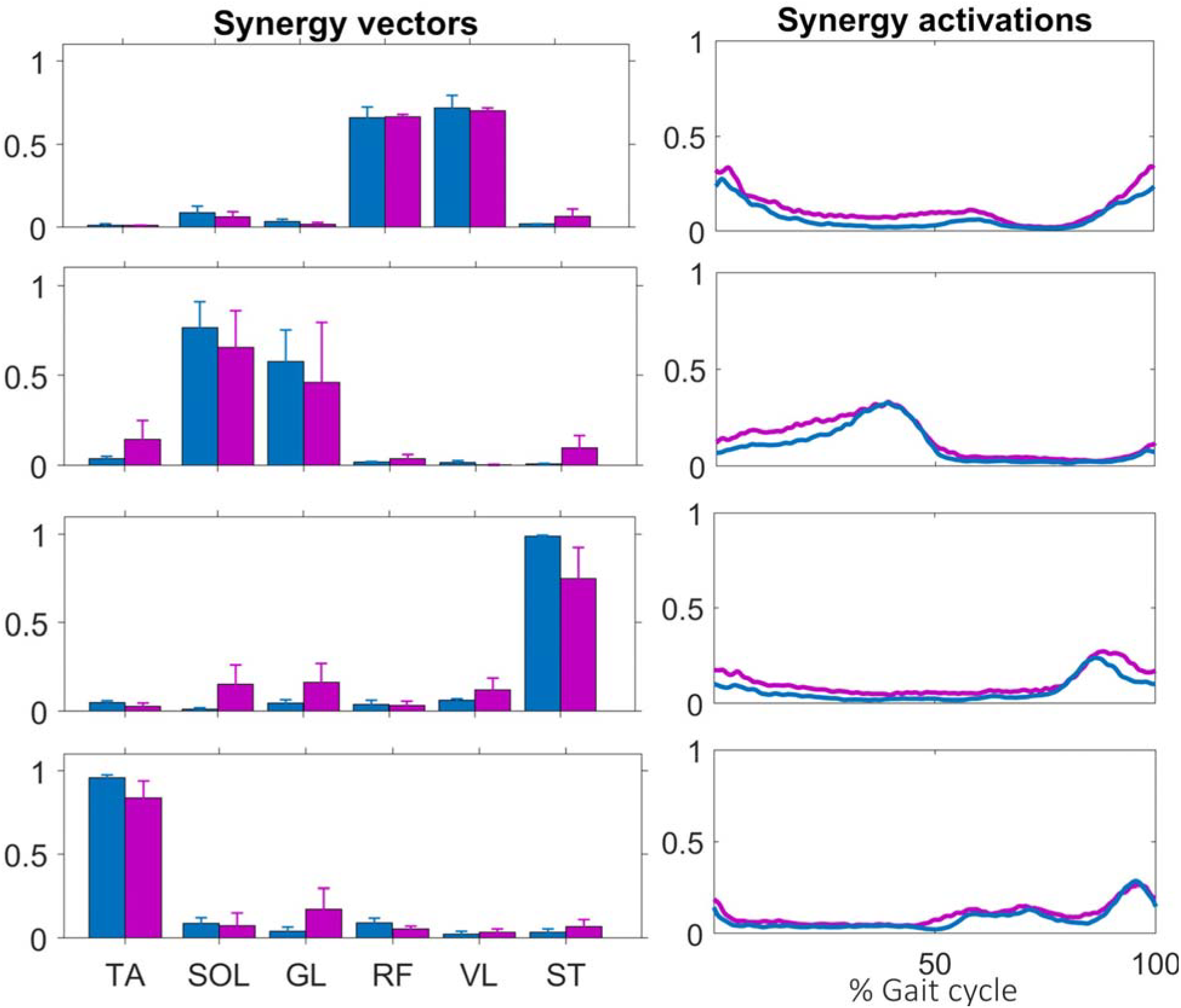
Reference muscle synergies extracted from control participants during the SW (blue) and the CW (purple) task. Muscle abbreviations: TA Tibialis Anterior, SOL Soleus, GL Lateral Gastrocnemius, RF Rectus Femoris, VL Vastus Lateralis, and ST Semitendinosus.

### 3.3. Muscle synergy analysis

On average, four muscle synergies were required to properly reconstruct the sEMG envelopes of control participants during both walking conditions (i.e., SW and CW), with an average VAF value of 93□±□2% during both SW and CW tasks, respectively. For individuals with CP, an increase in the number of muscle synergies (*p* < *0.05*) was observed during the CW task (n=4 [4-4]) compared to the SW task (n=3 [3-4]). An average VAF of 93%□±□1% and 92±□2% were obtained during SW and CW tasks, respectively.

Reference muscle synergies, obtained by averaging the muscle synergies of the control participants, are shown in **Figure 2. Figure 3** and **Figure 4** show the weight vectors (W) and the activation coefficients (C) of each CP patient during both walking tasks with the indication of the shared and specific muscle synergies.

**Figure 3.**
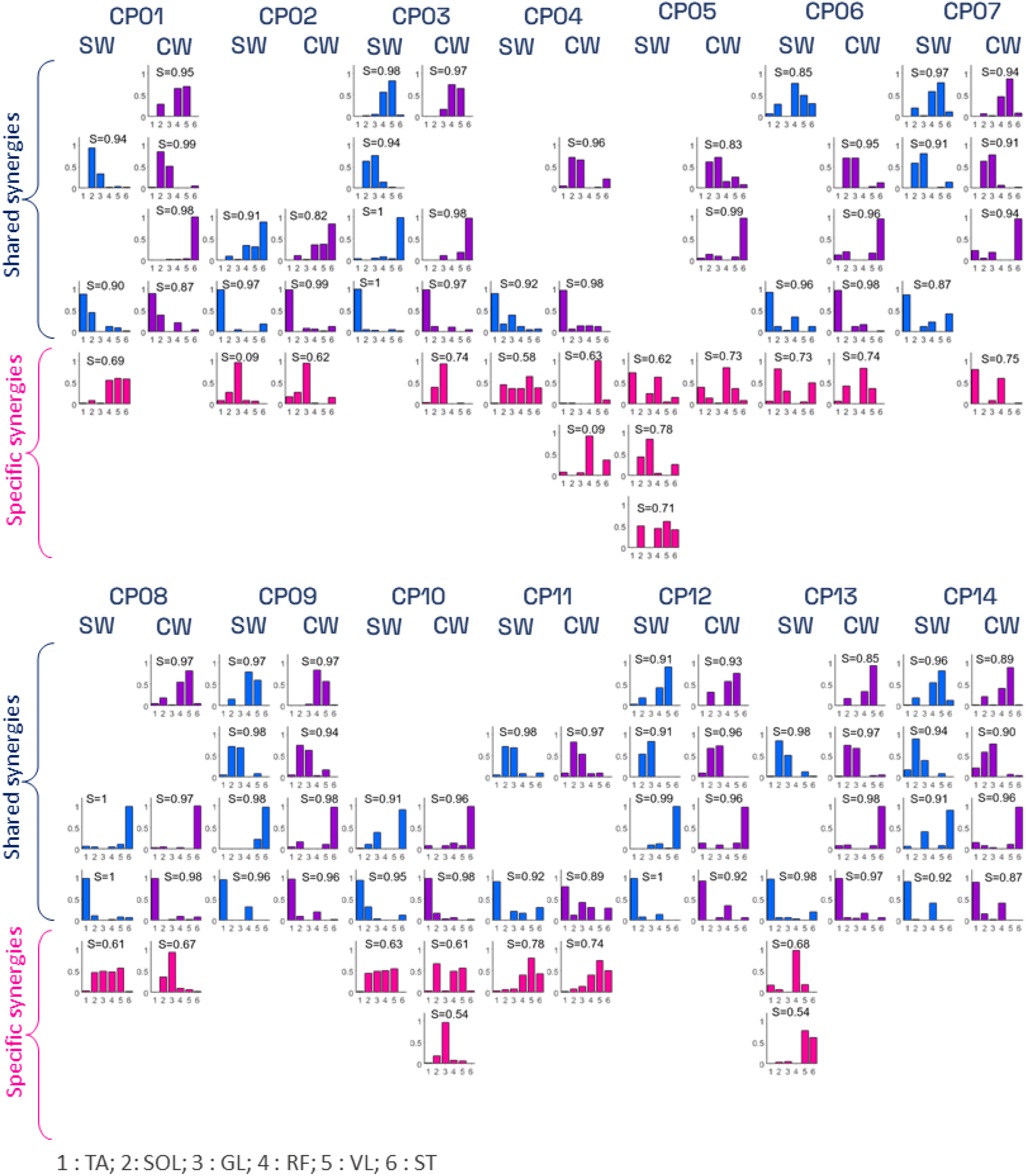
**Synergies’**weight vectors of CP participants during simple (SW) and complex (CW) tasks. Shared weight vectors are shown in blue and purple colors, while patient-specific weight vectors are shown in pink. For each weight vector, the cosine similarity (CS) value between the specific vector and the corresponding reference synergy is represented. Muscle abbreviations: TA Tibialis Anterior, SOL Soleus, GL Lateral Gastrocnemius, RF Rectus Femoris, VL Vastus Lateralis, and ST Semitendinosus.

**Figure 4.**
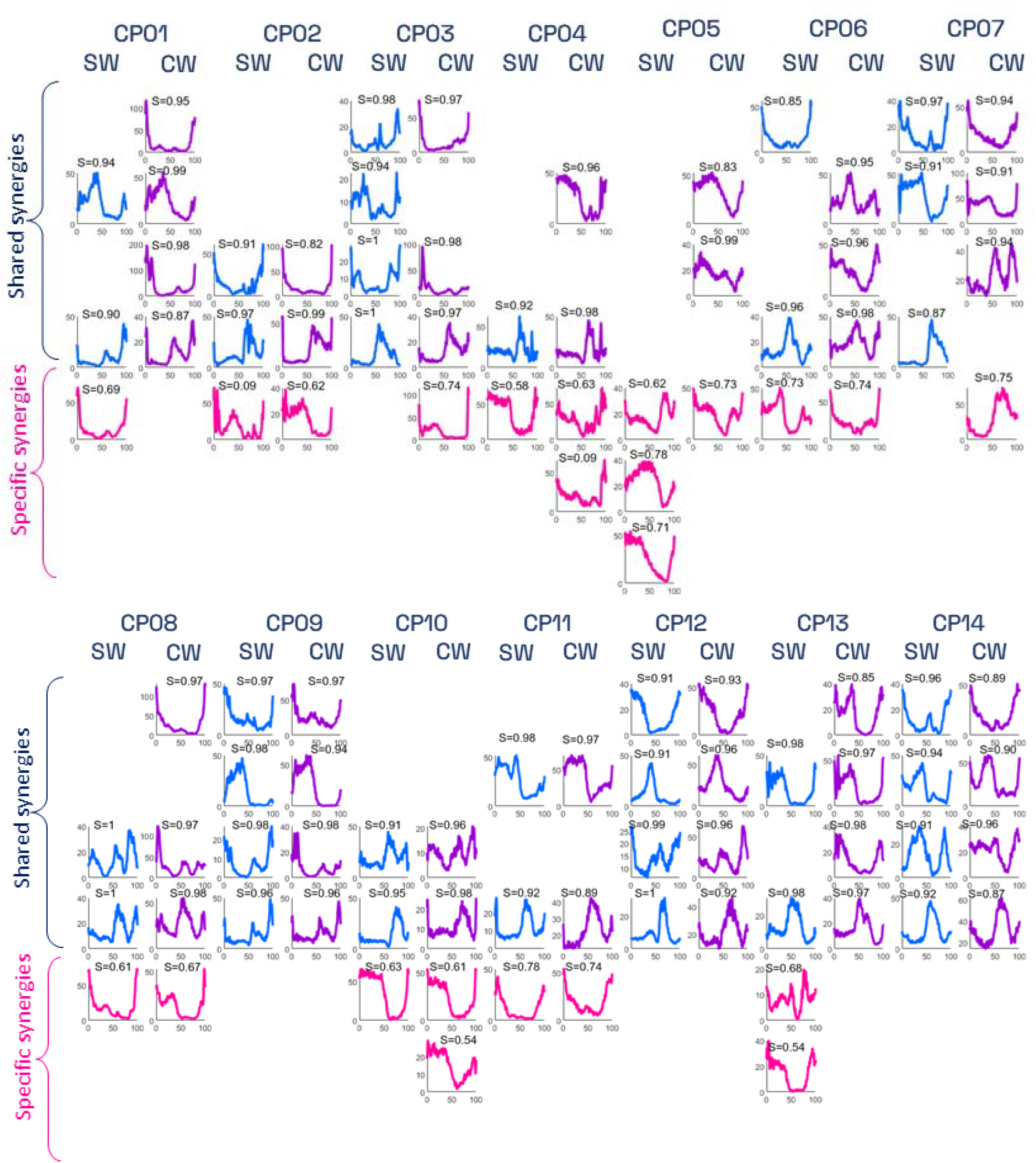
Synergies’ activation coefficients of CP participants during simple (SW) and complex (CW) tasks. Shared activation coefficients are shown in blue and purple colors, while patient-specific activation coefficients are shown in pink. For each activation coefficient, the cosine similarity (CS) value between the specific vector and the corresponding reference synergy is represented. Muscle abbreviations: TA Tibialis Anterior, SOL Soleus, GL Lateral Gastrocnemius, RF Rectus Femoris, VL Vastus Lateralis, and ST Semitendinosus.

Results showed the presence of *subject-specific* synergies (i.e., considered to be dissimilar to those of CTL participants) in 11 out of 14 CP participants. Only the three CP participants showing a GMFCS score of I (CP09, CP13, CP14) presented 4 *shared* muscle synergies (i.e., similar to those of CTL participants, with CS scores ranging from 0.87 to 0.99).

## 4. Discussion

The objective of this study was to assess the effect of a CW task on CST excitability and muscle synergies in individuals with and without CP. The two main findings are that: 1) compared to the SW task, the CW task induced a significant increase in corticospinal excitability and a modulation of muscle synergy structure in individuals with CP; and 2) both groups showed increased CST excitability during the CW task, no significant differences between groups were observed for this metric. This study is the first to investigate corticospinal excitability during walking in individuals with CP, providing essential information on the neuromuscular adaptations in this population.

### Corticospinal excitability

Partially consistent with our first hypothesis, both groups increased their CST excitability under the CW task. However, no differences in terms of MEP increase between groups were observed. Although the relationship between movement complexity and MEP size has not yet been established in the literature, our results align with several previous studies highlighting that complex motor tasks increase CST excitability in healthy subjects during manual (23,49,50) or locomotor tasks (34,51). For example, Barthélémy *et al.* (51) observed an increase in corticospinal excitability when participants walked against resistive forces. These results support the idea that CST excitability is modulated in a task-dependent manner, and may suggest that complex walking tasks can serve as powerful stimuli for motor system engagement. Moreover, it is acknowledged that MEP amplitude can be sensitive to cognitive processes such as precision demands(52) or decision-making (53). Thus, it is reasonable to speculate that heightened cognitive processing during the preparation of our visually guided task (*i.e.,* CW) may lead to an increase in CST excitability. Even if differences in MEPs amplitudes and shape were demonstrated between stroke survivors and healthy controls (54), our results revealed no differences in MEP area under the curve and increase between individuals with CP and controls. This is likely because the CP individuals included in this study were only mildly affected, as the majority have a GMFCS level of I or II. Then, the lack of group difference in MEP modulation may suggest that individuals with mild CP retain the capacity to modulate CST excitability in response to increased task demands, comparable to controls. However, the CP group showed a higher aMT than the control participants. This suggests that the higher stimulation intensity used relative to aMT in the CP group may have compensated for reduced corticospinal excitability, and then masking potential differences between the groups. Thus, similar MEP responses do not necessarily reflect equivalent CST reactivity (55), but may rather indicate reduced recruitment efficiency in CP, requiring greater stimulation to evoke comparable outputs. In conclusion, these preliminary results support that MEP may serve as valuable as a dynamic marker of CST function during functional tasks such as walking. It might be preferable to measure MEPs when people are moving rather than when they are at rest as this provides a better picture of their ability to function and adapt to their motor impairments.

### Muscle synergy modulation

Overall, individuals with CP recruited fewer muscle synergies than healthy controls during the SW task, indicating a general reduction in the complexity of their motor control system, which is often linked to poorer motor performance (9). In motor control systems with fewer muscle synergies, the central nervous system’s ability to independently control synergies responsible for different biomechanical functions is significantly compromised (14,45,56). The reduction in motor control system complexity can be observed through increased muscle coactivation, where weight vectors show the activation of a larger number of muscles, and decreased independence of neural control signals, indicated by activation coefficients that are more similar to each other and more overlapping throughout the task duration.

When moving from the SW task to the CW task, results demonstrated a reorganization in CP muscle synergies, in general revealing an increased number of muscle synergies, lower cosine similarity values compared to the reference muscle synergies, and an increased number of *subject-specific* muscle synergies. In other words, individuals with CP require a reorganization of their motor control strategies (in terms of both activation timing and muscle enrollment throughout the task duration) to cope with the new, more complex walking tasks. It aligns with studies suggesting that motor modules in individuals with neurological impairments can be modulated in response to environmental or biomechanical demands (57,58). CP group showed that either the spatial (i.e., weight vectors) or temporal (i.e., activation coefficients) components of the muscle synergies differed when compared to reference synergies of controls, who maintained highly consistent muscle synergies across walking conditions.

### Clinical implications

The findings of the current study highlight the potential for enhancing corticospinal excitability during a CW task. The results of this study showed a significant increase in CST excitability in both CP and control participants. These results suggest that individuals can increase their CST excitability to facilitate precise movement control in CW scenarios that demand accurate foot placement. Regarding muscle synergies, our results are in line with the literature highlighting that individuals with CP often employ simpler motor control strategies during gait compared to their typically developing peers (9). However, to obtain effective motor control during more complex locomotor tasks, individuals with CP required modulation of their muscle synergies by increasing motor control complexity (i.e., increasing the number of muscle synergies). Overall, the parallel increase in CST excitability and modulation of muscle synergies during the CW task in individual with CP suggest that task complexity may act as a powerful driver of neural and muscular adaptations. These results underline the value of implementing cognitively and sensorimotor challenging walking tasks in rehabilitation.

### Limitations

Some limitations of this study need to be highlighted. First, the sample size is relatively small sample size, which increases the risk of type 2 error and may impact the generalizability of the findings. Second, it is important to consider that the level of attention during the two walking tasks was not controlled, potentially affecting CST excitability. Third, the study did not include an analysis of the biomechanical changes and gait spatiotemporal parameters associated with the reduced number of muscle synergies in individuals with CP. Future studies should consider incorporating these elements to provide a more comprehensive understanding of the gait dynamics and functional implications in CP population.

## 5. Conclusion

This study has contributed to elucidating the mechanisms of gait control in CP. Additionally, our results clarified the increased involvement of the CST and the reorganization of motor control strategies in CP patients during more challenging walking tasks. These results pave the way for improved gait rehabilitation in CP by targeting complex tasks. Although our study measured CST excitability only in the TA muscle, future research should also investigate the SOL and RF muscles, as they play significant roles in walking.

## Data Availability

All data produced in the present study are available upon reasonable request to the authors

